# Systematic assessment of outcomes following a genetic diagnosis identified through a large-scale research study into developmental disorders

**DOI:** 10.1101/2020.10.23.20213157

**Authors:** H. Copeland, E. Kivuva, H.V. Firth, C.F. Wright

## Abstract

**Purpose:** The clinical and psychosocial outcomes associated with receiving a genetic diagnosis for developmental disorders are wide-ranging but under-studied. We sought to investigate outcomes from a subset of families who received a diagnosis through the Deciphering Developmental Disorders (DDD) study.

**Method:** Individuals recruited through the Peninsula Clinical Genetics Service who received a confirmed genetic diagnosis through the DDD study before August 2019 (n=112) were included in a clinical audit. Families with no identified clinical outcomes (n=16) were invited to participate in semi-structured telephone interviews.

**Results:** Disease-specific treatment was identified for seven probands (6%), while 48 probands (43%) were referred for further investigations or screening and 60 probands (54%) were recruited to further research. Just five families (4%) opted for prenatal testing in a subsequent pregnancy, reflecting the relatively advanced maternal age in our cohort, and 42 families (38%) were given disease-specific information or signposting to patient-specific resources such as support groups. Six interviews were performed (response rate=47%) and thematic analysis identified four major themes: reaching a diagnosis, emotional impact, family implications and practical issues.

**Conclusions:** Our data demonstrate that receiving a genetic diagnosis has substantial positive medical and psychosocial outcomes for the majority of patients and their families.

## Introduction

Developmental disorders describe a group of conditions that result in delayed cognitive and/or atypical physical development in children under 5 years of age ^1^. Whilst a wide range of aetiologies are recognised, up to 80% of cases are thought to be due to an underlying monogenic or chromosomal cause ^2,3^. Although standard clinical approaches are only able to identify diagnoses for less than half of affected individuals ^4^, family-based genome-wide sequencing studies have substantially increased the diagnostic yield over the last decade ^5,6^. The benefits of receiving a genetic diagnosis can be widespread, from counselling the family about their recurrence risk and offering pre-implantation/prenatal testing, through to identifying diagnosis-specific treatments for the proband ^7,8^. However, whilst a few recent studies have investigated the outcomes for families who received a clinical diagnosis through exome and genome sequencing, they mostly comprise case reports and small case series ^9^. To date, no research has systematically evaluated outcomes in a large cohort of patients who obtained their diagnosis through a research study.

The Deciphering Developmental Disorders (DDD) study ^4^ was created with the aim of using high-resolution genomic testing methods to investigate the genetic aetiology of abnormal development. From 2011-15, the study recruited ∼13,500 children with severe but previously undiagnosed developmental disorders, and their parents, from 23 National Health Service (NHS) Regional Genetics Services across the United Kingdom (UK) and one in Eire ^10,11^. Using a combination of high-resolution array-CGH testing and trio exome sequencing ^12,13^, the DDD study has identified plausible genetic diagnoses in around 40% of children and communicated these findings back to their local genetics teams for validation and discussion with the families ^11^. The DDD study thus provides an ideal cohort in which to systematically evaluate clinical outcomes arising from a paediatric genomics research study.

Here we describe a pilot study investigating the clinical and psychosocial outcomes of receiving a genetic diagnosis in a subset of 112 DDD participants from a single UK centre. Our study will not only help better understand the wider impact of receiving a diagnosis for the patient and their family, but also inform larger systematic outcomes studies.

## Methods

We used a mixed methods approach to gather outcomes data on DDD families, which combined a quantitative clinical audit and a series of semi-structured qualitative interviews. Eligibility for inclusion in the audit was defined as any individual recruited to the DDD study through the Peninsula Clinical Genetics Service (which covers Devon and Cornwall) for whom the DDD study had identified a definite genetic diagnosis (Figure 1). Eligible individuals were identified through the DECIPHER database ^14^, and linked-anonymised

**Figure 1.**
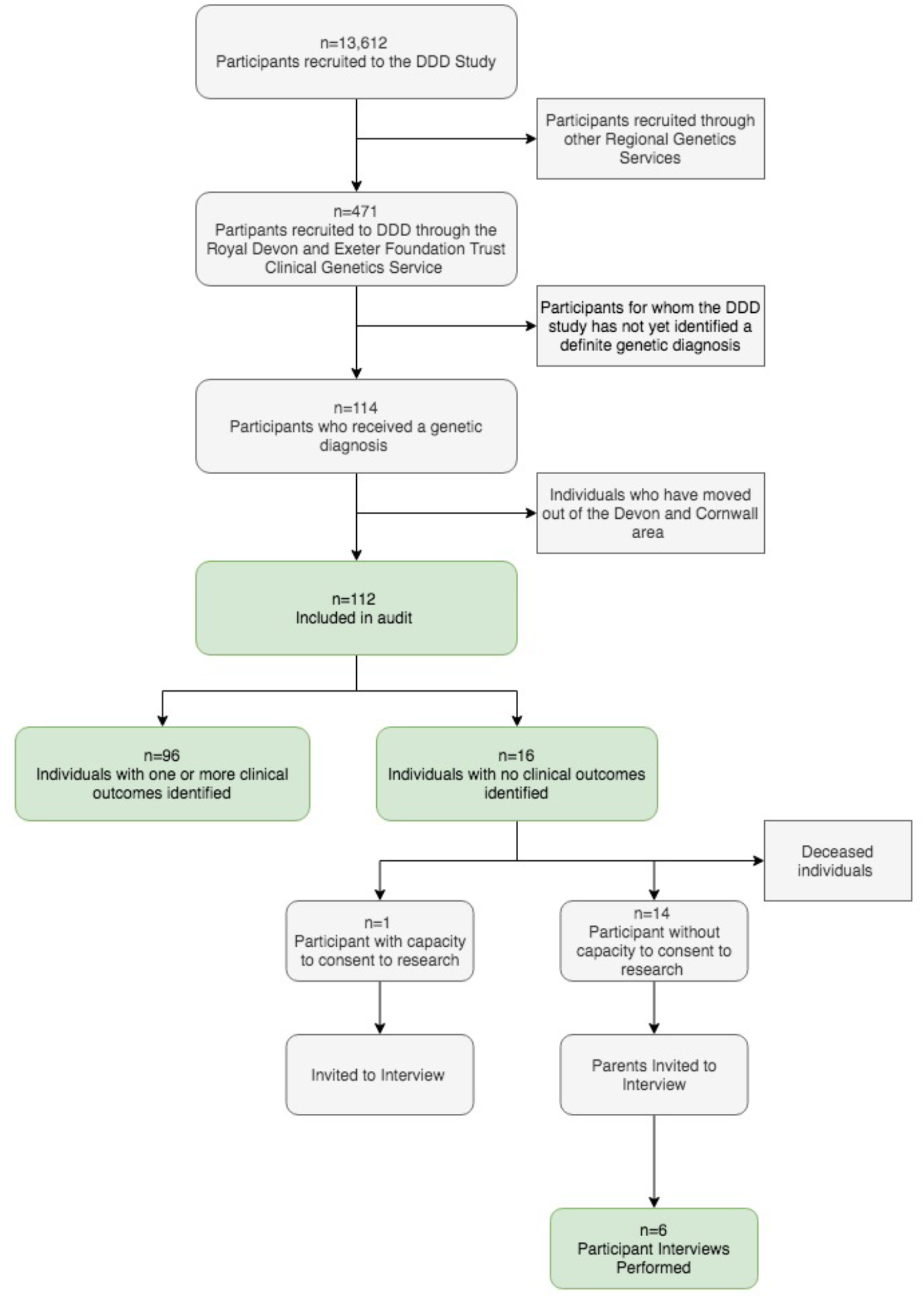
Flowchart showing recruitment process for the clinical audit and the semi-structured interviews.

DECIPHER IDs were used by the local team to connect the genetic diagnosis with the local NHS Clinical Genetics records for each proband. We developed a pro forma for collecting standardised information during the clinical audit relating to outcomes, including treatments, further investigations, reproductive decisions, adverse events, recruitment to further genetic research studies, and the information given to the patient/parents at the time of diagnosis (Supplementary Table 1). Disease-specific treatments were identified through review of correspondence in clinical records and through published literature searching. We also used the Unique patient support website ^15^ and other online resources to identify whether gene-disease-specific information was available, such as a leaflet or document written for patients and their families.

Following the clinical audit, families in whom we were unable to identify any outcomes were invited to participate in a semi-structured interview. We designed a questionnaire (Supplementary Table 2) to identify personal, family and wider outcomes that were not possible to assess through the audit. Interviews were performed over the telephone and recorded to enable transcription. A deductive thematic approach was used to analyse the qualitative data to identify major and minor themes.

## Results

We identified 112 diagnosed probands that were eligible for inclusion in the clinical audit (Figure 1). The probands ranged in age at recruitment from 3 to 44 years (median=8.7) and the age of the parents at the birth of the proband ranged from 17 to 66 (median=31), which is representative of the rest of the DDD cohort (median=31, p=0.3). A range of confirmed diagnoses were identified, including 97 dominant (of which 81 were *de novo*), 12 recessive and 3 X-linked recessive. Within the cohort, variants were classed as pathogenic or likely pathogenic following the ACGS best practice guidelines for variant classification ^16^ and validated in an accredited laboratory. Diagnostic variants were identified in 85 different genes, with diagnoses in 16 genes being causative in more than one unrelated individual.

Clinical outcomes identified in the audit (or subsequent interview) are summarised in Figure 2. Seven probands (6%) received a diagnosis for which a disease-specific treatment was known (Table 1). Of these, 5/7 probands had treatable epilepsy as part of their phenotype, where knowledge of the specific molecular cause allows the correct drug to be rapidly selected from those available ^17^. One proband received an expedited hematopoietic stem cell transplant to treat bone marrow failure, and another may receive treatment for Parkinsonism when age appropriate.

**Table 1.** Summary of diagnoses where specific treatments were available.

| DECIPHER ID | Gene | Treatment | Primary Phenotype Treated | Uptake | Comment |
| --- | --- | --- | --- | --- | --- |
| 275863 | *ATP1A3* | Benzodiazepines, dopamine agonists or levo-dopa | Epilepsy | Y | Treatment changes reported to reduce frequency and severity of seizures |
| 305456 | *GRIN2B* | N-methyl D-aspartate receptor blockers such as Memantine or Radiprodil | Epilepsy | N | Patient's seizures already reasonably controlled with different medication |
| 282007 | *KCNQ2* | Carbamazepine | Epilepsy | Y | Patient was prescribed carbamazepine which was reported to reduce seizure frequency |
| 273914 | *PHGDH* | L-serine | Epilepsy | Y | Treatment did not have a significant impact on patient's symptoms. Patient was aged 4 years when treatment given; highest efficacy rates reported when given *in utero* |
| 264089 | *SCN2A* | Lamotrigine | Epilepsy | Y | Treatment changes reported to improve seizure control |
| 291232 | *LIG4* | Hematopoietic stem cell transplantation | Bone marrow failure | Y | Prophylactic hematopoietic stem cell transplant performed 1.5 years after the diagnosis |
| 275418 | *WDR45* | Parkinson treatments | Early parkinsonism (teens) | N | Patient was not yet in their teens |

**Figure 2:**
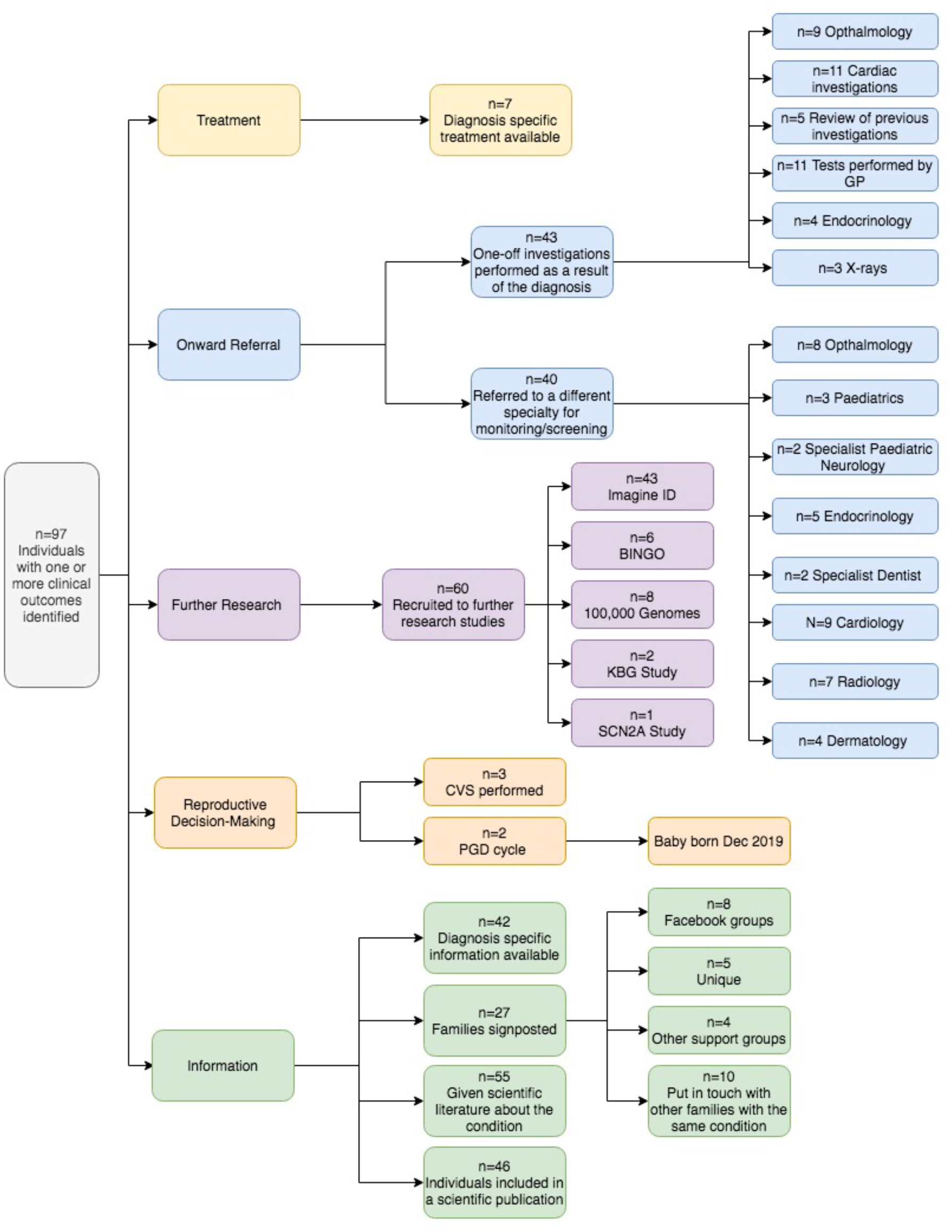
Flowchart demonstrating outcomes identified primarily through the clinical audit.

A total of 43 further diagnostic clinical investigations and 40 referrals to different medical specialties were requested for 48 individuals (43%). These included requests for reviews of previous scans (n=5), ophthalmology assessments (n=9) and cardiac investigations (n=11), and referrals for ongoing screening by radiology (n=7), ophthalmology (n=8) and endocrinology (n=5) departments. Many of these individuals did not present with problems requiring those medical specialties pre-diagnosis. Furthermore, we were able to identify two participants for whom the diagnosis prevented unnecessary diagnostic interventions, including growth investigations and an MRI scan under general anaesthetic.

A total of 60 families (53%) were recruited to additional research studies following their diagnosis, including the Intellectual Disability and Mental Health: Assessing the Genomic Impact on Neurodevelopment (IMAGINE ID) study ^18^ (n=43) and the Brain and Behaviour in Intellectual Disability of known Genetic Origin (BINGO) study ^19^ (n=6). Gene or disease-specific information in the form of a patient information leaflet or document was given to 42 families (37.5%). A total of 27 families (24%) were signposted to sources of additional information, including the Unique website (n=5), Facebook groups (n=8) and disease-specific support groups (n=4). Contact between the families of probands with the same diagnosis was facilitated for 10 families.

Discussions with parents regarding reproductive choices were documented in the notes of 45 families, which represents 83% of couples where the mother was of reproductive age (<45 years) at the point of receiving the proband’s diagnosis. The audit identified three cases in which the proband’s diagnosis was used to direct invasive prenatal testing in subsequent pregnancies, all of which were negative. In a further two cases, the couples chose to have pre-implantation genetic diagnosis; treatment for one couple is still ongoing, and the second couple now have a healthy child. These five families represent 9% of the families in which the mother was of reproductive age.

The audit identified one adverse effect of receiving a diagnosis. The proband showed distress during their results appointment upon learning that the diagnosis would likely lead to additional symptoms and screening in the future, requiring further face-to-face and telephone consultations to manage their distress and anxiety.

We were unable to identify any outcomes beyond the diagnosis itself for 16/112 individuals in the clinical audit (Figure 1). Of these, one proband was deceased, so 15 families were invited to participate in a telephone interview. Seven initial responses were received (47% response rate) from parents of patients, and six interviews were conducted. The probands in these six families had all received a diagnosis of a *de novo* variant identified through the DDD study. Thematic analysis identified four major themes and a range of subthemes, shown in Figure 3, and selected quotes from interview transcripts are given in Table 2.

**Table 2.** Selected quotes from semi-structured interviews with six DDD parents, demonstrating the identified themes.

|  |  |  |
| --- | --- | --- |
| Reaching a Diagnosis | **Motivations for joining the DDD Study** | *"I think anybody who has a child with additional needs feels a need to know"*<br>*"[...] and I would say the added hope was that if someone else had a child with the same problems as [child] then they would have more information earlier"* |
|  | **Diagnostic challenges** | *"Because she's never fit into any specific categories, there was nothing the doctors could put their fingers on [...] as parents it was incredibly frustrating"*<br>*"I think having [...] I call it the false diagnosis, I remember sitting there thinking [...] this isn't [child], because I'd obviously researched [condition] and I always felt that [child] didn't fit the picture and I tried desperately to make him fit, like really desperately"* |
|  | **Feelings about involvement in the study** | *"I just find the whole thing so interesting and I'm pleased that we took part"*<br>*"It's made a huge difference to me as a mum, me as a parent and me as a person, having that knowledge is a great thing, and it wouldn't have happened without the project"* |
| Practical Impact | **Day-to-day support** | *"[...] it has always been a point of frustration for me that I have needed to write [child's] symptoms in a long list over and over again, and so it feels nice to just write the diagnosis and be done with it"* |
|  | **Support networks** | *"[child] has always been compared to normal children her whole life [...] it would be nice to be able to compare her to others with the same situation for a change"*<br>*"The helpful bit is knowing the information, it's action rather than talking about it"* |
|  | **Educational support** | *"She was put into mainstream secondary school [...] that first year, it destroyed her, it absolutely destroyed her [...] the diagnosis came in and now she's in a special school [...] she's now a lot happier, she's back on track, she's flying so I can't be happier [...] if the diagnosis hadn't come through I'm honestly not sure of where she'd be"* |
| Emotional Issues | **Guilt** | *"I think it would be true to say that any parent, particularly a mother [...] has a massive burden of guilt [...] it seemed impossible to get away from the sense that I could have done something more about it, done less [...]. It's really such a life-changing feeling to know that nothing I did or could have done has affected the way she is"* |
|  | **Relief** | *"It helped [child] a huge amount because it's given her credibility, and that, for me as her dad, is a huge relief"*<br>*"I don't think either of us would have blamed the other one [...] it was just a relief to know that we hadn't affected her life in any way, because it's always in the back of your mind"* |
|  | **Uncertainty** | *"There seems to be a limited amount of information about [condition] life expectancy for example [...] which is always a massive concern to parents, you know, are they likely to outlive us?"*<br>*"[...] we're very much aware that [child] has an uncertain future, [...] the diagnosis can't predict his future so there is no path to follow [...] I think we've addressed those issues about his uncertain future already"* |
| Family Impact | **Family Members** | *"[...] at least now I can turn around to my girls and say it's not hereditary [...] it doesn't mean that your children are going to be disabled [...] I can't describe how valuable that feels"* |
|  | **Future Pregnancies** | *"It was a very worrying time for me, I had all of the tests and everything and they all came back fine [...] but the worry was still there, and I honestly didn't enjoy the pregnancy. It felt like a cycle of being anxious and stressed, but then feeling guilty because I knew stress could cause problems and that cycle didn't really go away [...] I had buried those feelings over the years [...] I only really remembered when my son's wife was pregnant and it was like history repeating itself again"* |

**Figure 3.**
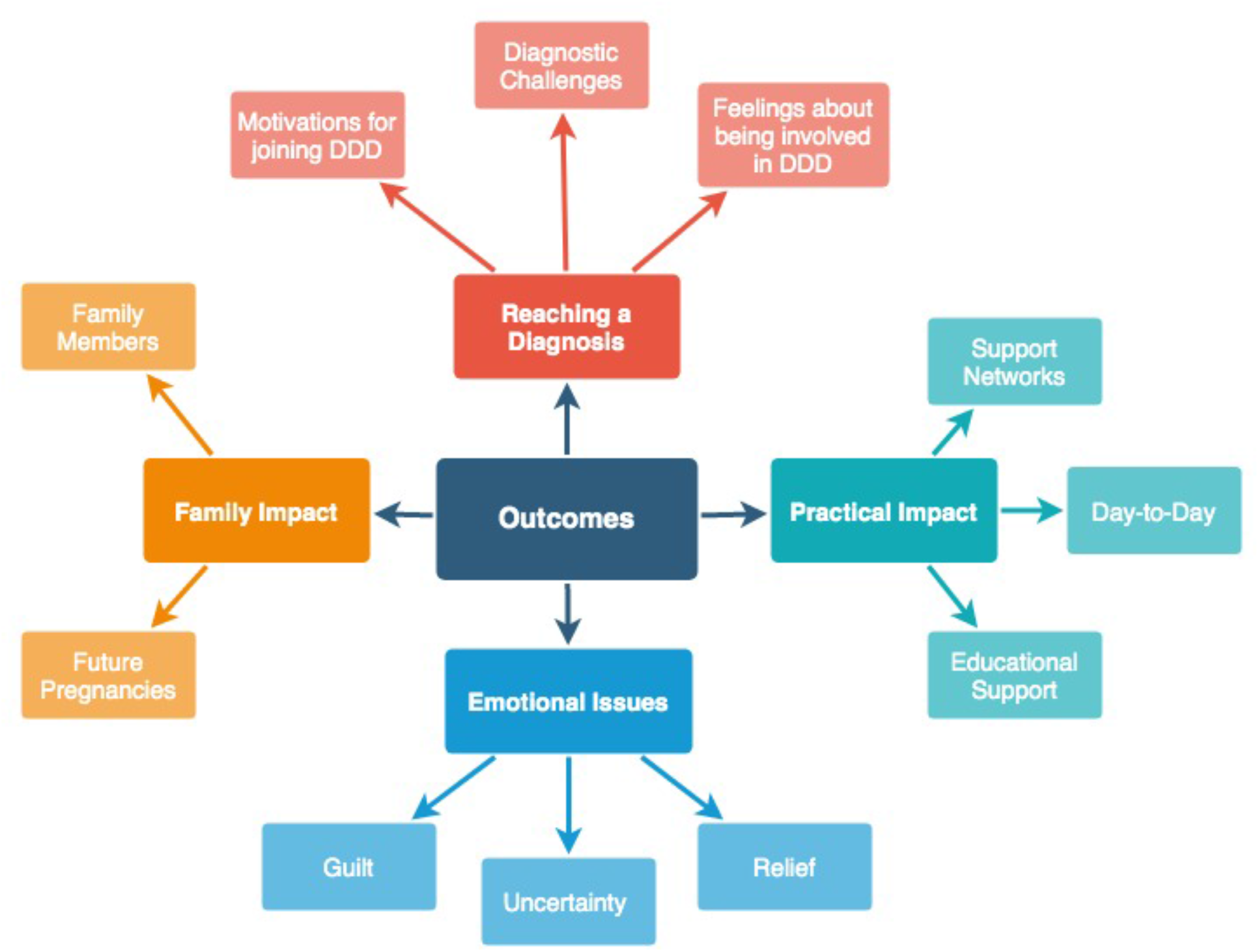
Cluster diagram representing the four major themes and their respective sub-themes identified through thematic analysis of the interview transcript data.

All interview participants were positive about having participated in the DDD study, and one participant described a treatment change that had arisen for their child following the diagnosis that was not identified in the clinical audit (included in Figure 2 and Table 1). The main motivation for participating in the study was finding an answer for their child’s problems, with many describing a long and complicated search for a diagnosis ^20^. Parents felt a sense of responsibility to obtain a diagnosis, and also described secondary altruistic motivations. Two families described the negative impact of previous incorrect diagnoses, with one interview participant reporting difficulty in accepting a diagnosis that didn’t seem to fit the child’s problems. Most participants reported feelings of guilt about their child’s difficulties, and upon learning that the genetic variant has arisen *de novo*, some participants reported that their guilt had reduced. Whilst all participants acknowledged that their child’s diagnosis had provided clarity as to the cause of their symptoms, for some, the diagnosis also brought uncertainty, particularly amongst parents of younger probands (n=3 with children <15 years).

All participants discussed the impact of their child’s diagnosis on other family members, and valued the relevance of the information for the wider family. Three participants raised the topic of further children and provided different perspectives of the decision-making process faced by families without a diagnosis. One participant described a subsequent pregnancy before the final diagnosis as an extremely challenging time; the lack of clarity about the cause of her child’s symptoms negatively impacting her experience of the subsequent pregnancy. Another participant described how not having a diagnosis previously had impacted on the couple’s decision not to have another child.

Despite feeling generally positive about receiving a diagnosis, all six interviewees explained that the level of day-to-day needs of, and support for, their child had not substantially changed following the diagnosis. Nonetheless, parents did feel that the process of applying for support had been streamlined by having a definitive genetic diagnosis rather than a list of symptoms. One participant described the diagnosis as being key to accessing specialist educational support, which significantly improved the child’s wellbeing. There was disparity among the interview participants about accessing support networks; interestingly, those who were aware of similarly affected children often did not wish to make contact due to fear of overwhelming the other family or because they did not perceive a benefit to such a meeting.

## Discussion

We have evaluated the clinical and psychosocial outcomes in a subset of 112 families from a single UK centre who received a genetic diagnosis from the DDD study. We were able to identify diagnosis-specific outcomes in 97/112 (87%) individuals, including direct medical outcomes for the proband in 45% of cases, such as diagnosis-specific treatments (n=7) and referral for additional investigations or screening (n=43). Those patients who received a diagnosis-specific treatment experienced clinical improvements in symptom control, demonstrating the value and importance of tailored treatments ^21,22^, and emphasising the need for continued research into this field. In contrast to treatment of existing phenotypes, referral for additional screening often related to monitoring and managing phenotypes that had not yet emerged, something which caused anxiety in at least once case.

Some families (n=5) were able to access prenatal testing and pre-implantation genetic diagnosis treatments, which would not have been available to them without the diagnosis. We anticipate that many more would have accessed this option had the parents been of reproductive age at the point of receiving the diagnosis. Accurate genetic diagnosis of developmental disorders empowers individuals to make informed and value-based reproductive decisions, highlighting the importance of a diagnosis for family planning purposes ^23,24^. For many participants (53%), receiving a diagnosis meant they met the eligibility criteria for further research studies and subsequent successful recruitment suggests that they had a positive perception of genetic research following their involvement in the DDD study. The audit identified 46 participants who had been included in a scientific publication, demonstrating the benefits of novel diagnoses to the scientific community. Additionally, we found that around a third of participants were given patient-focussed information relating to their diagnosis whilst others were signposted to sources of additional information; both are ways to reinforce complex genetic information. In 55 cases, where patient-focussed information was not available, scientific literature was instead given to the family where appropriate. These findings highlight and support the need for continued development of patient-focussed information to be developed as new developmental disorders are identified.

Our semi-structured interviews focused on families in whom no clinically relevant outcomes could be identified from the clinical audit. Although this strategy left us with just 16 eligible families, we judged that it would maximise our chances of identifying additional outcomes whilst biasing the participants towards those who might feel less favourably towards genetics research. Nonetheless, the high response rate of nearly 50% amongst those invited to interview suggests high engagement and motivation within the group, despite no outcomes having been identified through the clinical audit. Moreover, through the course of just six interviews, we identified an additional family in whom diagnosis enabled a specific treatment. This suggests that we under-ascertained outcomes based on a clinical audit of genetics notes and indicates that further research using extended hospital notes or additional interviews is needed to fully explore outcomes.

All participants reported feeling positively about their involvement in the DDD study, and whilst recognising the potential for bias, this finding supports the value for patients and families of being involved in genetics research. Four main themes and further subthemes were identified following thematic analysis, although there is significant overlap between the themes (Figure 3). The observed sense of parental responsibility to find an explanation for their child’s symptoms links to a reported sense of failure when standard clinical pathways fail to find an answer ^25^. Secondary altruistic motivations demonstrate the different expectations families have of participating in genetic research studies ^26^. Experiences shared by interview participants about the diagnostic odyssey emphasised the complex interplay between their child’s phenotypic symptoms, creating an emotional burden ^20^. One family described their experience with earlier misdiagnoses, demonstrating the widespread negative impact on the patient and the wider family of incorrect diagnoses. This further reflects a theme of parental “gut instinct” ^27^; some parents reported feeling strongly that a diagnosis was incorrect, but did not feel qualified to question or challenge the medical opinion ^28^, potentially increasing self-doubt and further delaying a correct diagnosis ^29^.

Most interview participants described feeling significant levels of guilt about their child’s problems. For them, finally knowing the underlying cause changed this guilt, demonstrating the value of timely diagnosis ^30,31^. However, this outcome is partly influenced by the fact that all interviewed families had a diagnostic *de novo* variant; higher levels of parental guilt have been linked to inherited autosomal recessive or X-linked diagnoses in children ^30^. Relief was a frequently reported emotional impact associated with receiving a diagnosis. Participants associated relief with feelings of validation for their child’s difficulties and their own concerns. Relief has also been linked to parental empowerment ^32^, showing the interplay between parental guilt and relief, and highlighting the value of finally obtaining a diagnosis. Concern for other family members also demonstrates that the impact of not having a diagnosis extends beyond the individual patient, and reflects the importance of a family-based approach in managing highly penetrant monogenic conditions ^33^. Being able to accurately clarify and communicate risks to family members was highly valued by the interview participants, with emotional responses reflecting a sense of parental responsibility in providing information for the whole family ^25^. Interview participants differed in their decision to have another child and their uptake of testing during subsequent pregnancies, highlighting the complexities of such decisions. The interviews demonstrate that these emotions can last for a number of years, and can resurface in the future, emphasising the emotional burden felt by parents without a diagnosis. One participant’s reflection that having an earlier diagnosis might have influenced the decision to have another child emphasises the challenges faced in reproductive decision-making in a rapidly changing field.

Our findings are limited by a few practicalities. Firstly, although genetic and phenotypic data on DDD families is managed centrally and shared with clinical teams via the DECIPHER database, a linked-anonymised system of IDs is used and researchers do not have access centrally to personal identifiers, which are maintained inside local NHS clinics. We therefore limited our sample group to one UK Regional Genetics Service, so our conclusions may not be representative of different services or geographies. Secondly, medical records at the Royal Devon and Exeter NHS Foundation Trust are not currently electronic, and clinical notes from different specialties are often held in numerous different locations and institutions across the region. Our clinical audit was limited to notes held within the Peninsula Clinical Genetics Service, so actions undertaken by other specialties may have been missed. Thirdly, we decided to focus our interviews on individuals in whom we were unable to identify any outcomes from the clinical audit, so our sample size for the qualitative phase of the study was small. Future research could utilise the same methodology with larger sample sizes across multiple genetics services (led by individuals embedded within each Regional Genetics Service to access local patient notes), or could explore similar themes in additional participants regardless of whether a clinical outcome had been recorded in the notes.

Knowledge of the underlying molecular aetiology of disease has intrinsic value and is a worthwhile outcome of genetic testing and research ^34,35^. Nonetheless, showing how that knowledge is used and to what, if any, clinical purposes it can be put is of great importance for assessing the wider medical, social and economic significance of a genetic diagnosis. This study aimed to systematically assess both clinical and non-clinical outcomes associated with receiving a diagnosis through a subset of participants in the DDD study. We show that receiving a diagnosis contributes to clinically relevant outcomes in a high proportion of individuals with developmental disorders and can streamline future clinical care. Although a disease-specific treatment was available to only a small number of individuals at the time of writing, the majority of these treatments were associated with an improvement in symptoms and more targeted treatments are likely to become available in future. Furthermore, through the interview process, we found that even for those individuals without clinical outcomes, participating in a genetics research study was a positive experience and receiving a diagnosis can have significant personal, emotional and psychosocial outcomes for the patient and their wider family.

## Data Availability

The authors confirm that the data supporting the findings of this study are available within the article and its supplementary materials.

## Acknowledgement

The authors would like to thank the patients and families involved in the study. The authors would also like to thank Profs Matt Hurles, David Fitzpatrick and Mike Parker for their helpful input, in addition to Drs Carole Brewer, Peter Turnpenny, Julia Rankin, Claire Turner, Bruce Castle, Charles Shaw-Smith and Emma Baple for their support. The DDD study presents independent research commissioned by the Health Innovation Challenge Fund [grant number HICF-1009-003], a parallel funding partnership between the Wellcome Trust and the Department of Health, and the Wellcome Sanger Institute [grant number WT098051]. The views expressed in this publication are those of the author(s) and not necessarily those of the Wellcome Trust or the Department of Health. The study has UK Research Ethics Committee approval (10/H0305/83, granted by the Cambridge South REC, and GEN/284/12 granted by the Republic of Ireland REC). The research team acknowledges the support of the National Institute for Health Research, through the Comprehensive Clinical Research Network. This study uses DECIPHER (https://decipher.sanger.ac.uk), which is funded by the Wellcome Trust. This work was originally submitted as part of an MSc thesis in Genomic Counselling at the University of Manchester.

---

**Supplementary Table 1.**
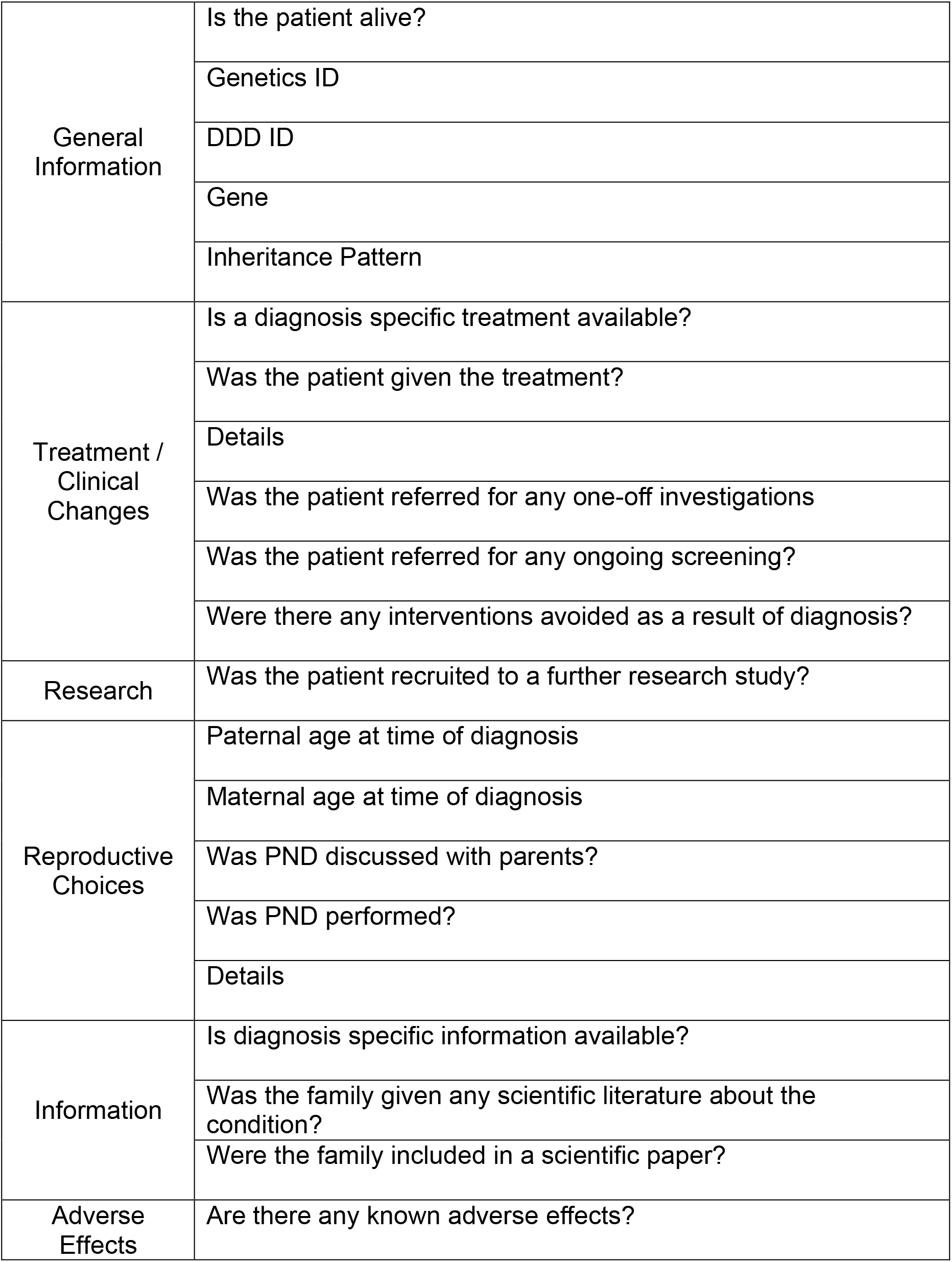
Pro forma used to identify outcomes through the clinical audit from the genetics notes of participants

**Supplementary Table 2.**
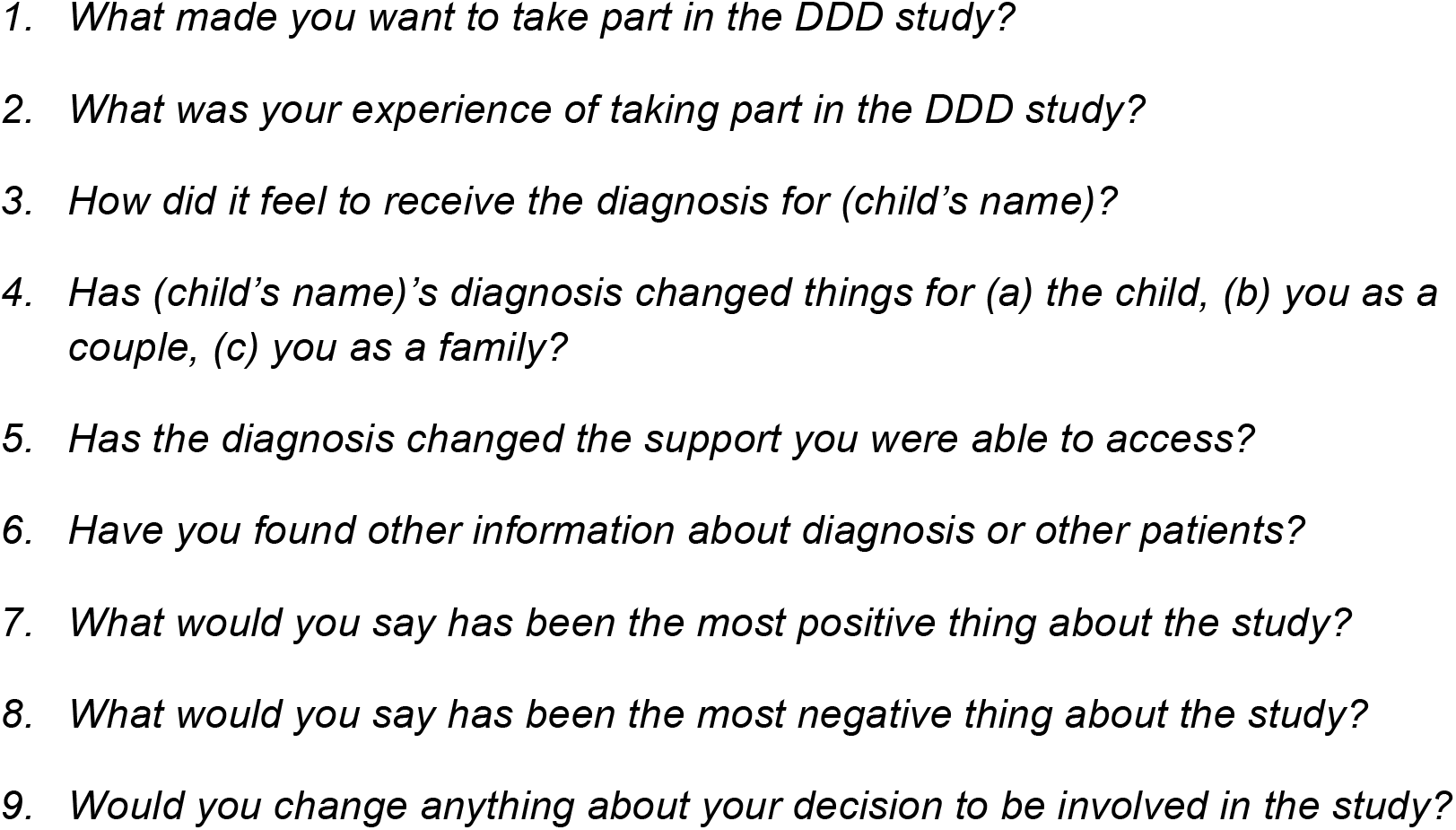
Semi-structured Interview Questions used to evaluate non-clinical outcomes

